# SLaM Image Bank – a real-world diverse London cohort linking brain MRI to electronic mental health and dementia records for the development of clinical decision support tools using artificial intelligence

**DOI:** 10.1101/2025.01.07.25319950

**Authors:** Ashwin V. Venkataraman, Paul Wright, Amelia Jewell, Christopher Webb, Yini Zhang, Soha K. Patil, M. Jorge Cardoso, James H. Cole, Matthew J. Kempton, Charles R. Marshall, Mitul A. Mehta, Christoph Mueller, Timothy R. Nicholson, Thomas A. Pollak, Timothy Rittman, Zeynep Sahin, Ana B. Solana, Daniel R. Stahl, František Váša, Allan H Young, Steve Williams, Robert Stewart, Dag Aarsland

**Author notes:** corresponding author Dr Ashwin V. Venkataraman (corresponding author), Department of Old Age Psychiatry, IoPPN, 6th Floor, M6.01, Box P070, De Crespigny Park, Denmark Hill, London, SE5 8AF.

## Abstract

**Purpose:** Rapid developments are occurring in artificial intelligence (AI) and machine learning (ML) applied to neuroimaging. To date, advances in this space have largely been limited to research cohorts with little real-world translation that is clinically meaningful for patients in psychiatry and neurology and those with associated neuropsychiatric symptoms. There is a lack of large real-world multimodal linked datasets combining MRI imaging, clinical variables and other biomarkers within more representative ethnically diverse and deprived populations with multiple neuropsychiatric and systemic co-morbidities, alongside the right infrastructure to link them.

**Participants:** We linked brain MRI scans in South London and Maudsley (SLaM) NHS Trust (UK), with clinical data from electronic mental health and dementia records for patients across multiple clinical sites in South London harnessing the Clinical Record Interactive Search (CRIS) data platform from 2008-2022.

**Findings to date:** 12,547 patients (age range 6–108, female 54%, male 46%) were identified with 14649 unique MR studies totalling 58620 MRI scans across 5 scanners with linked clinical data, from an ethnically diverse population (44% non-White), most with high social deprivation (n = 7424), and having a variety of diagnoses, of which F00-F09 Organic was the largest ICD category (n = 4628, 33% of total), followed by F20-F29 Schizophrenia and related disorders (n= 2151, 15% of total), and F30-F39 Mood [affective] disorders (n = 1607, 12% of total). The commonest recorded diagnoses were Alzheimer’s Disease (n = 2849), Schizophrenia (n = 1021) and mild cognitive impairment (n = 740).

**Future Plans:** SLaM Image Bank is an exceptionally rich real-world cohort with linked MRI and clinical data reflecting the diversity of the population of South London. It provides a unique platform for future testing of automated decision support tools that may identify unique signatures of diagnosis and neuropsychiatric symptom subtype, markers of prognosis, and provide stratification for interventions or trials that are potentially clinically meaningful. This will form the basis to which new linked data and MRI data will be added in order to grow this cohort alongside the potential additions of other imaging modalities, novel biomarkers, meta-data and patient groups in the future.

**Strengths and limitations:**

- SLaM Image Bank is a diverse large brain image cohort that grows with new scans linking MRI data to electronic mental health and dementia records from South London and Maudsley NHS Trust in London, UK, with data from 12,547 patients with a variety of psychiatric/neurological diagnoses over a 13-year period.
- The aim is to provide clinically meaningful translation with an initial focus on identifying unique signatures of disease subtype and neuropsychiatric symptoms, markers of prognosis, and stratification for interventions or trials in a move towards real-world precision medicine alongside epidemiological and mechanistic research opportunities.
- This is an imaging cohort that accrues from pre-existing data and does not require additional and expensive recruitment exercises which may in turn lead to a powerful, translational and scalable framework to build upon.

## Introduction

Harnessing brain imaging for the benefit of patients with psychiatric and neurological illnesses has the potential to provide signatures of disease that are clinically meaningful [1,2]. There are rapid developments occurring in artificial intelligence (AI) and machine learning (ML) across many fields in medicine, including psychiatry and neurology. In particular, AI/ML and predictive modelling coupled to neuroimaging has the ability to detect previously unseen features that may be relevant to diagnosis and prognosis, and to stratification for treatment. Currently, MRI images in a clinical approach are typically only visually described or rated using a variety of scores by radiologists depending on the underlying aetiology [3–5]. The highly dimensional nature of neuroimaging data in the form of 3D voxel information, high degrees of covariance within the data, and that voxel intensities are non-linear mean that neuroimaging as a modality is particularly suited to AI/ML applications, which may in turn assist in the drive towards precision medicine [6,7]. Despite widespread research these AI/ML approaches have not translated into clinical practice [8].

To date, translation of clinically meaningful decision support tools has been limited by various barriers. At the forefront of these are the lack of rich clinically meaningful data from real-world healthcare settings often limited by different research protocols. The majority of data generated using AI/ML applied to neuroimaging in patients with psychiatric and neurological conditions have been from large research data resources such as ENIGMA or ADNI [9–11], but there are various biases (e.g. age, ethnicity, level of deprivation) that have limited the generalisability and scalability of these to real-world settings[12]. UK Biobank, whilst exceptionally powerful in its size and depth of information, also lacks representativeness as a volunteer sample, and does not adequately capture diverse patient groups[13]. Few, if any, AI/ML models on big data have been trained on neuroimaging data from real-world settings specifically from those less represented individuals with ethnic and socio-economic diversity, co/multimorbid pathologies, and with a higher burden of co-existing neuropsychiatric symptoms [1]. This is particularly timely given recent advances in AI allowing harmonisation of heterogeneous clinical MRI scans to leverage research grade information [14]. This is especially important given the potential of future disease modifying therapies in certain diseases such as Alzheimer’s disease [15–19] alongside the delivery of primary and secondary prevention of modifiable risk factors [16,20–22] and the shift in conceptualisation of earlier life psychiatric illness into similar research paradigms.

We therefore established the South London and Maudsley (SLaM) Image Bank, drawing on a large real-world diverse cohort of people in psychiatric and dementia care services, linking MRI to routinely collected clinical data and pre-existing meta-data. The primary aim of SLaM Image Bank is to create a platform for testing automated decision support tools based on neuroimaging and associated features that may identify unique signatures of disease and neuropsychiatric symptom subtypes, provide prognostic information, allow stratification of subgroups in ways that are precise, clinically meaningful, and also of benefit to interventions/trials. New prospective data will be iteratively added to this cohort with UK Biobank scan sequence alignment to allow wider comparison. SLaM Image Bank has been set up initially for memory and brain health clinics[23] provided by the South London and Maudsley NHS Trust in the UK (see below) and recently expanded to all clinical sites in the Trust. We believe that the model has the potential for wider implementation to support a multi-site network of self-accruing cohorts drawing on routine data in wider patient groups. In addition, we are harnessing the strengths of the Clinical Record Interactive Search (CRIS) (see below) [24] to leverage natural language processing (NLP) extraction of meta-data from both clinical and neuroradiology report text and to draw on established linkages with external health and social care data, as well as to incorporate other imaging modalities and additional patient cohorts in future.

## Cohort description

### Setting

The South London and Maudsley (SLaM) National Health Service (NHS) Trust is the largest mental health provider in Europe, with a long history of mental health provision dating back to the Bethlem Hospital in 1247. It sits within the NHS, a public healthcare system that is free of charge. Dementia assessment and care is largely provided by mental health trusts in the UK and included here. It covers a catchment population of over 1.3 million people across the four London boroughs of Croydon, Lambeth, Lewisham and Southwark. Descriptive data from these populations are available here [25]. In this population minority ethnic groups form substantially higher proportions compared to England. This geographic catchment area also includes those from higher areas of deprivation as measured by the Index of Multiple Deprivation (IMD), which is a comprehensive measure used to analyse socioeconomic status in specific geographic areas combining various indicators across economic, social, and housing factors such as income, employment, health, education, housing, crime, and living environment and automatically calculated based on a patient’s post code.
The Clinical Records Interactive Search (CRIS) was developed in 2007-08 in order to provide researcher access to SLaM’s electronic Patient Journey System (ePJS), thus allowing in-depth secondary analysis of naturalistic healthcare data. It allows extraction of numerical, string and free text data, with additional NLP capability built in over the past 10 years, whilst simultaneously preserving anonymity through technical and procedural safeguards[25]. It includes clinical outcome data on patients sometimes many years after MRI, making it also suitable for prognostic modelling.

### SLaM Cohort

The SLaM Image Bank cohort is currently a retrospective cohort of 12,547 patients who have been under a clinical service in SLaM NHS Trust from January 1^st^ 2008 to January 1^st^ 2022 with the availability of 13,950 linked clinical brain MRI scans including 1403 repeat scans. SLaM catchment provision is currently structured within the following specialty groupings: Addictions; Behavioural and Developmental Psychiatry; Child and Adolescent Mental Health Services; Mental Health of Older Adults and
Dementia; Mood, Anxiety and Personality; Psychological Medicine and Psychosis, with associated similar academic groupings at King’s College London. Ethnic groups were either Asian/Asian British, Black/Black British, other ethnic groups, White or unknown. Hospitalisation, mortality data, social care and functional measures are being acquired. This project is approved under CRIS Ethics (Oxford REC, reference 18/SC/0372), sub-project ID CRIS 21-123 and part of the NIHR BRC Maudsley Neuroimaging Call (NQOD-04). Consent and anonymisation follow CRIS opt out protocols as described in [24,26].

### Linkage and data architecture

An overview of the SLaM Image Bank linkage technical architecture is shown in Figure 1 below.

The SLaM Image Bank linkage first identified patients according to above criteria via CRIS where data is stored in a SLaM Firewall/Safe Haven. The SLaM clinical data linkage comprises one part of the King’s Health Partners Academic Health Sciences Centre (KHP AHSC) which was established with King’s College London, Guy’s and St Thomas’ and King’s College Hospitals NHS Foundation Trusts. Full details on CRIS, data extraction, storage, processing, de-identification, and NLP are detailed here [24]. MRI Images are stored and processed on the KCL neuroimaging analysis network with high performance computing (HPC) cluster processing capability for de-identified linked images.

### Initial data curation

Data were organized according to the Brain Imaging Data Structure (BIDS)[27] by categorising the type of scan (BIDS suffix) and use of contrast according to the DICOM series description attribute. Localizer and calibration scans were excluded, as were any that indicated a non-brain body part (e.g. “spine” or “orbits”).

## Findings to date

### Study population

Of the 12547 available MRI scans from patients and matched clinical data, 6764 (54%) were female, and 5783 (46%) were male with an age range of 6 to 108 with no missing data in these categories. Ethnic groups were subdivided into larger categories with the highest proportion of patients being White (British/Irish/White-other) (56%, compared to 60% in London, and 86% nationally), Black/Black British (African/Caribbean/Black-other) (25%, compared to 13% in London, and 3% nationally), and Asian (Indian/Pakistani/Bangladeshi/Chinese/Asian-other) (8%, compared to 18% in London and 8% nationally). 4% were not known. (Table 1 and Figure 2A)

**Table 1.**
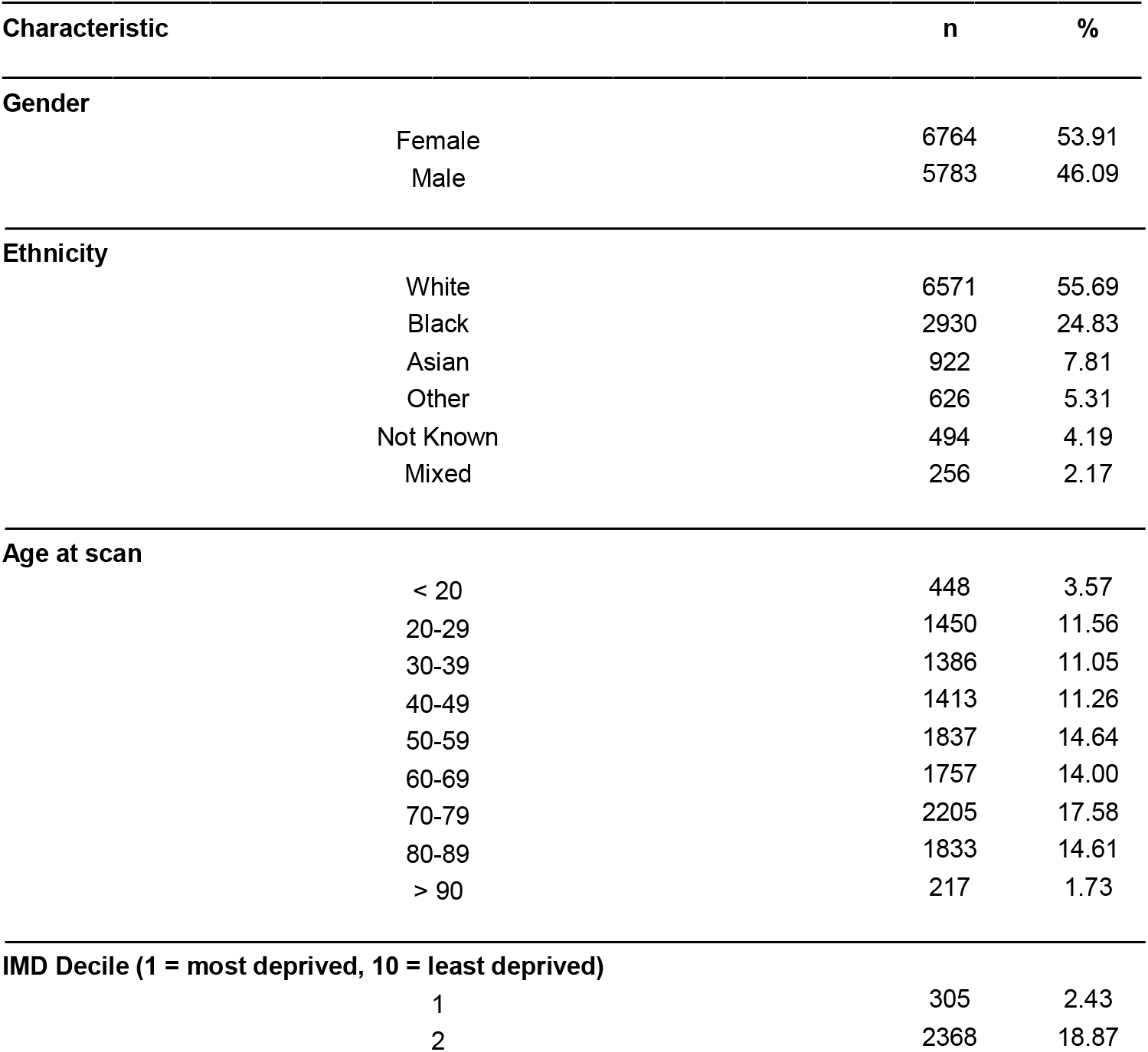
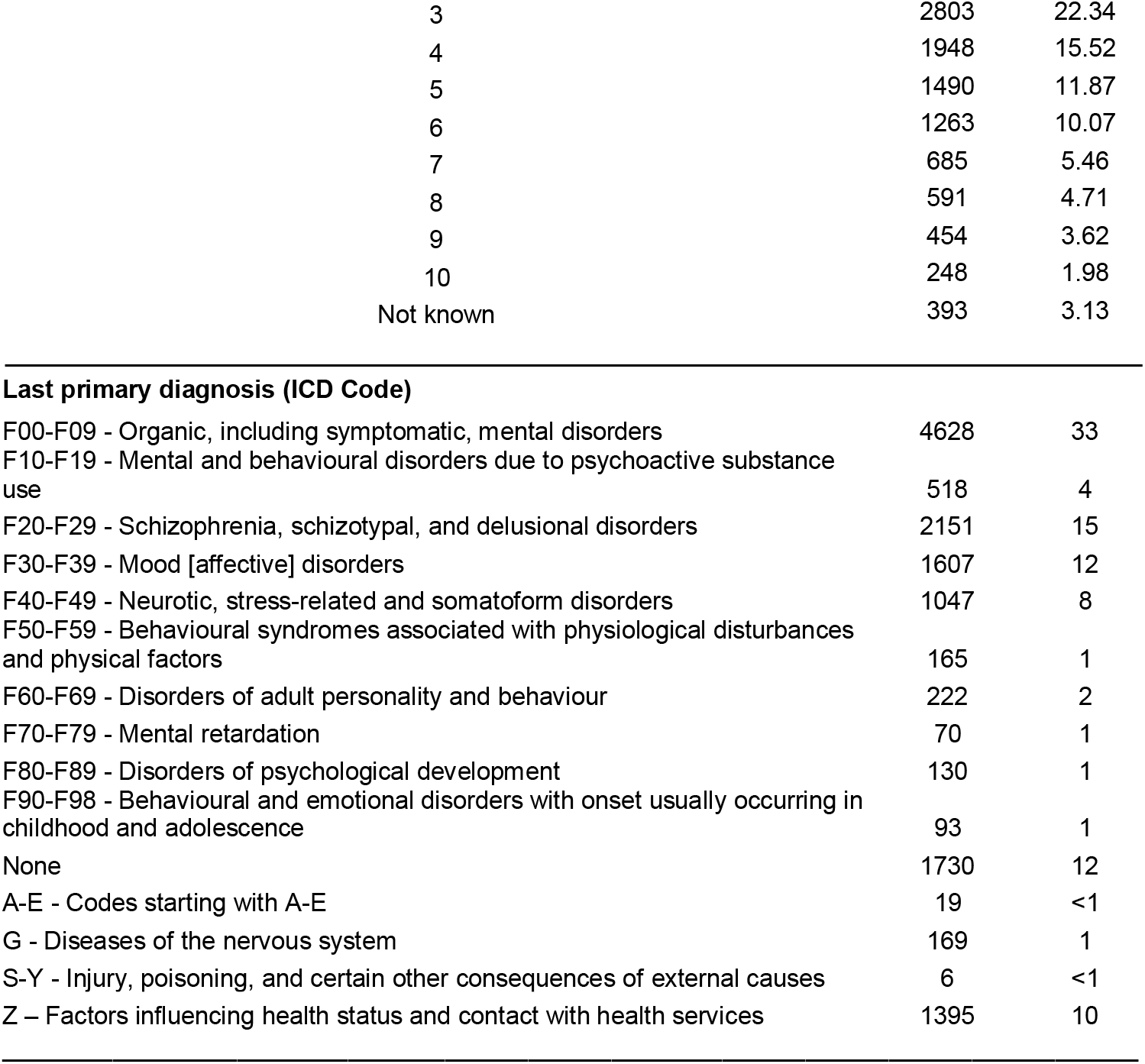
Descriptive statistics of the 12,547 patients to date in the SLaM Image Bank

| Characteristic | n | % |
| --- | --- | --- |
| **Gender** |  |  |
| Female | 6764 | 53.91 |
| Male | 5783 | 46.09 |
| **Ethnicity** |  |  |
| White | 6571 | 55.69 |
| Black | 2930 | 24.83 |
| Asian | 922 | 7.81 |
| Other | 626 | 5.31 |
| Not Known | 494 | 4.19 |
| Mixed | 256 | 2.17 |
| **Age at scan** |  |  |
| < 20 | 448 | 3.57 |
| 20-29 | 1450 | 11.56 |
| 30-39 | 1386 | 11.05 |
| 40-49 | 1413 | 11.26 |
| 50-59 | 1837 | 14.64 |
| 60-69 | 1757 | 14.00 |
| 70-79 | 2205 | 17.58 |
| 80-89 | 1833 | 14.61 |
| > 90 | 217 | 1.73 |
| **IMD Decile (1 = most deprived, 10 = least deprived)** |  |  |
| 1 | 305 | 2.43 |
| 2 | 2368 | 18.87 |
| 3 | 2803 | 22.34 |
| 4 | 1948 | 15.52 |
| 5 | 1490 | 11.87 |
| 6 | 1263 | 10.07 |
| 7 | 685 | 5.46 |
| 8 | 591 | 4.71 |
| 9 | 454 | 3.62 |
| 10 | 248 | 1.98 |
| Not known | 393 | 3.13 |
| **Last primary diagnosis (ICD Code)** |  |  |
| F00-F09 - Organic, including symptomatic, mental disorders | 4628 | 33 |
| F10-F19 - Mental and behavioural disorders due to psychoactive substance use | 518 | 4 |
| F20-F29 - Schizophrenia, schizotypal, and delusional disorders | 2151 | 15 |
| F30-F39 - Mood [affective] disorders | 1607 | 12 |
| F40-F49 - Neurotic, stress-related and somatoform disorders | 1047 | 8 |
| F50-F59 - Behavioural syndromes associated with physiological disturbances and physical factors | 165 | 1 |
| F60-F69 - Disorders of adult personality and behaviour | 222 | 2 |
| F70-F79 - Mental retardation | 70 | 1 |
| F80-F89 - Disorders of psychological development | 130 | 1 |
| F90-F98 - Behavioural and emotional disorders with onset usually occurring in childhood and adolescence | 93 | 1 |
| None | 1730 | 12 |
| A-E - Codes starting with A-E | 19 | <1 |
| G - Diseases of the nervous system | 169 | 1 |
| S-Y - Injury, poisoning, and certain other consequences of external causes | 6 | <1 |
| Z – Factors influencing health status and contact with health services | 1395 | 10 |

**Figure 1.**
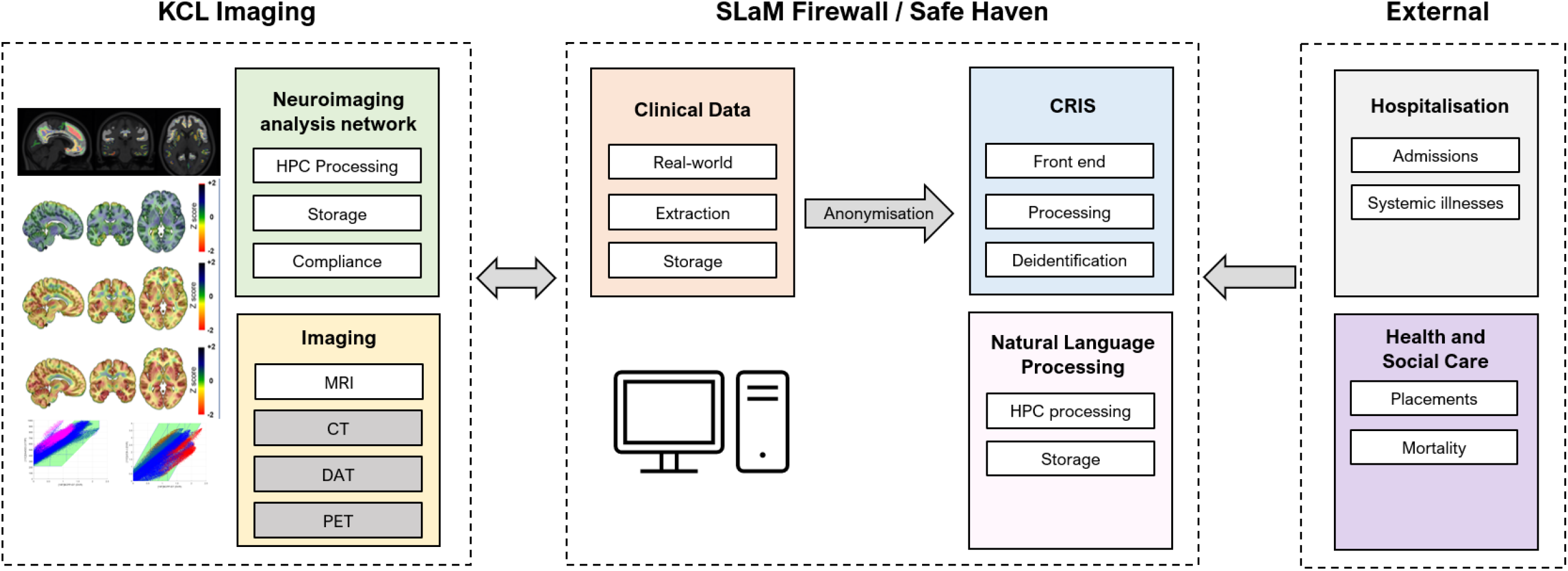
Diagram of SLaM Image Bank linkage technical architecture including KCL Imaging with the Neuroimaging Analysis Network and imaging modalities, SLaM Firewall/Safe Haven with clinical, CRIS and NLP Processing, and links to external hospitalisation and health and social care data sources. White boxes indicate established linkages, and grey are included in future plans.

**Figure 2.**
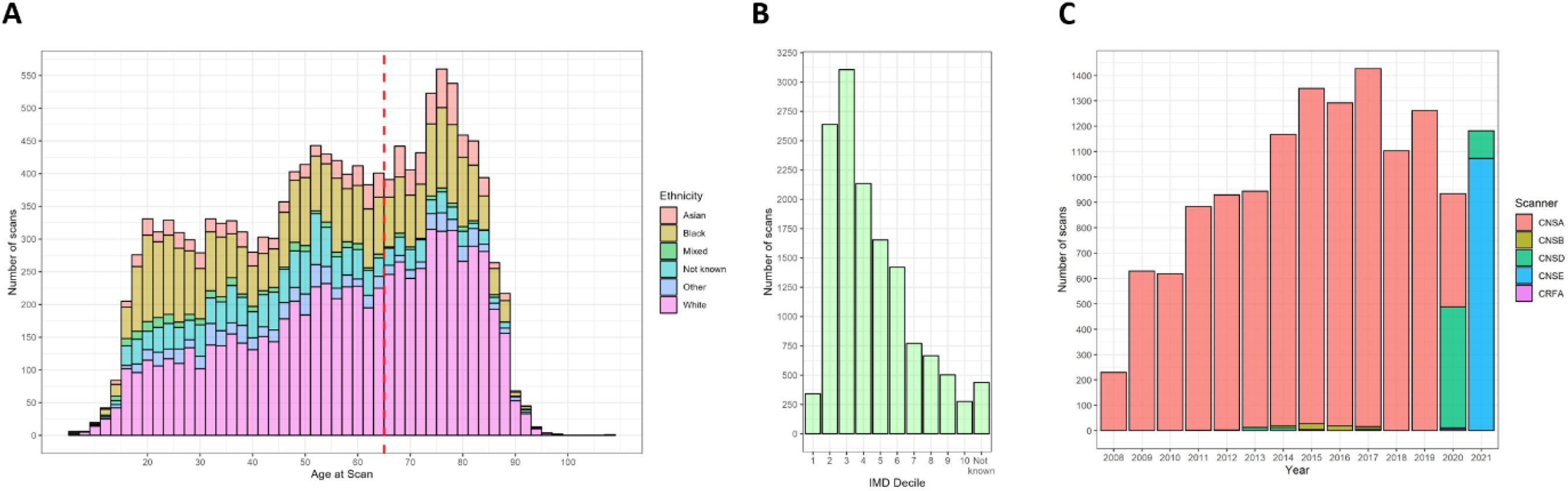
A) Stacked histogram showing the number of patients with MRI scans and linked clinical records stratified by age and ethnicity, with the red dashed line at 65 representing a common age cut-off to distinguish general adult and older adult services. B) Bar chart of the number of patients with MRI scans stratified by IMD Decile where 1 is most deprived, and 10 is least deprived. C) Bar chart of the number of MRI scans acquired per year stratified by scanner type (CNSA GE 1.5T HDx, CNSB GE 3.0T HDx, CNSD GE 3.0T MR750, CNSE GE 1.5T Signa Artist, and the CRFA GE 3.0T MR750).

Diagnoses were extracted pre and post MRI scan, and at subsequent available time intervals post scan coded according to ICD-10 classification [28]. The last primary diagnoses following an MRI scan are detailed in Table 1 and stratified according to age in Figure 2A, with the latest diagnosis derived from individual ICD codes shown in Figure 3A and 3B.

**Figure 3.**
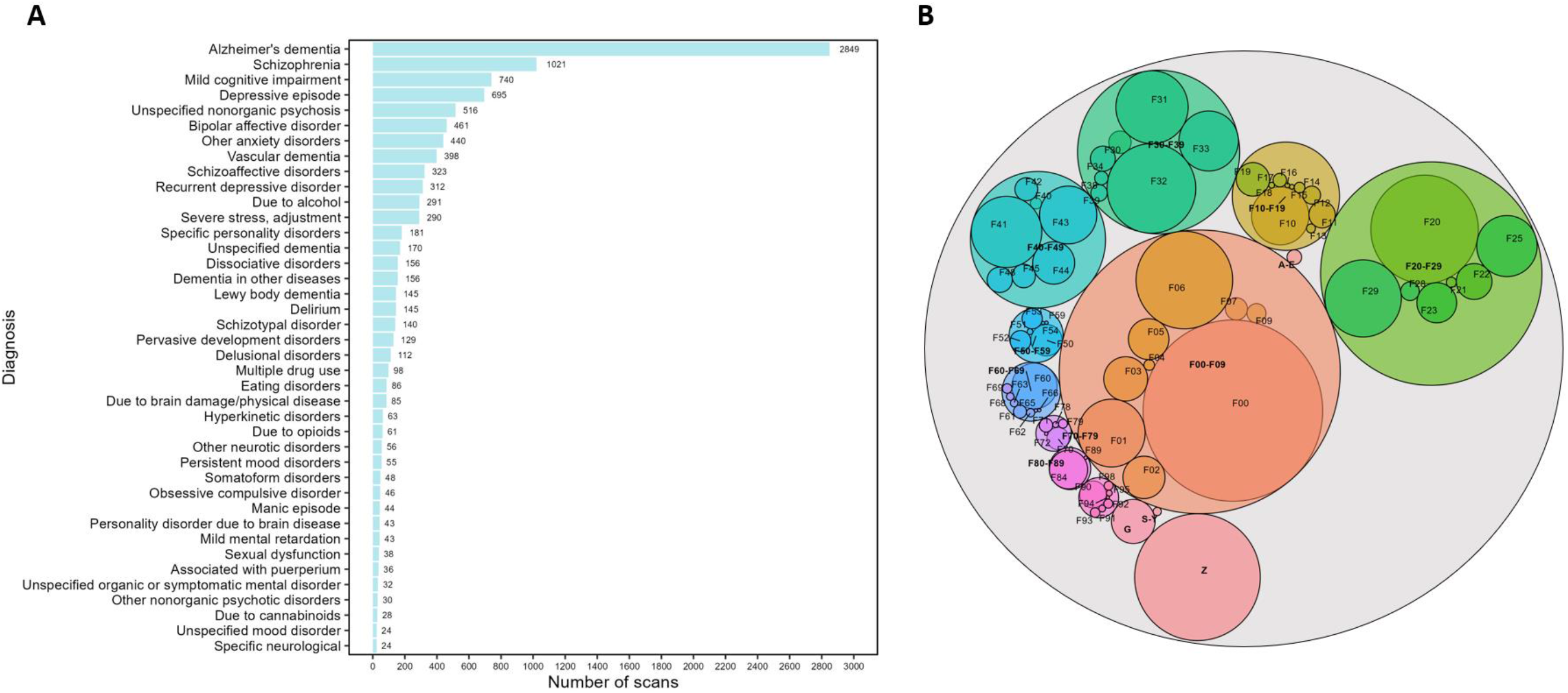
A) Bar chart showing the latest clinical diagnosis (ICD10 level to one decimal point) post scan following MRI acquisition derived from ICD categories stratified according to number of scans where diagnoses below 20 scans and those with no diagnosis are not shown. B) Proportional bubble chart for all ICD10 codes where the size corresponds to the number of scans available and the larger categories are in bold.

The majority of patients with an MRI scan and linked clinical record were from memory services or old age community mental health teams which is reflected in the last primary diagnoses recorded with 4628 scans (33% of the total cohort) within ICD codes F00-F09 – Organic, including symptomatic mental disorders (Table 1, Figure 2A). Of this category the majority of scans (n = 2849) were in patients with Alzheimer’s disease (Figure 3A) in keeping with national memory clinic data [29].

The second largest diagnostic category was F20-F29 Schizophrenia, schizotypal and delusional disorders with 2151 scans (15% of the total cohort) made up primarily of MRI scans from early intervention psychosis teams and community mental health teams. Within this category the majority of scans (n = 1021) were in patients with schizophrenia.

Other notably large diagnostic categories included F30-F39 Mood disorders with 1607 scans (12% of the total cohort), and F40-49 Neurotic, stress related and somatoform disorders having 1047 scans (8% of the total cohort).

The IMD scores stratified by decile are shown in Table 1 and Figure 2B at the time of the MRI scans with 1 being most deprived, and 10 being least deprived. There is a clear positive skew with 2803 scans from the 3^rd^ decile (22%), 2368 scans from the 2^nd^ decile (19%), and 1948 scans from the 4^th^ decile (16%). IMD scores above 10% are over-represented in SLaM.

### Comorbidities

There is a pressing need to understand co-morbidities in neuropsychiatric disease, particularly to understand their implications on patient outcomes and treatment strategies, alongside brain specific effects [21,30]. We classified all common co-morbidities (21 conditions extracted via MedCAT from CRIS) in this imaging cohort using NLP extraction and stratified according to < 65 or ≥ 65 years old. In those < 65 years old the most common comorbidity was falls (37%), followed by diabetes (29%), hypertensive disorder (26%), hypertension (21%) and asthma (16%). In patients ≥ 65 years old the most common comorbidities were falls (71%), hypertensive disorder (62%), hypertension (47%), diabetes (42%) and stroke (23%). All extracted comorbidities are shown in Figure 4.

### Imaging

During this study period 14649 unique MR studies and corresponding clinical neuroradiology reports were acquired from patients. These were acquired across 5 different scanner models – CNSA General Electric 1.5T HDx, CNSB General Electric 3.0T HDx, CNSD General Electric 3.0T MR750, CNSE General Electric 1.5T Signa Artist, and the CRFA General Electric 3.0T MR750. See Figure 2C for the breakdown of number of scans sessions of each type stratified by year.

Depending on the scan indication, various imaging protocols were used. The majority of patients have T1-weighted structural MRI (3D MPRAGE), T2-weighted structural MRI (2D Axial T2, 2D Axial FLAIR), and depending on patient compliance and scanner type there would also be additional scans including, but not limited to, cerebral blood flow measures (3D Arterial Spin Labelling for Perfusion), 3D Susceptibility Weighted Imaging and at least 40-direction diffuse tensor imaging.

**Figure 4.**
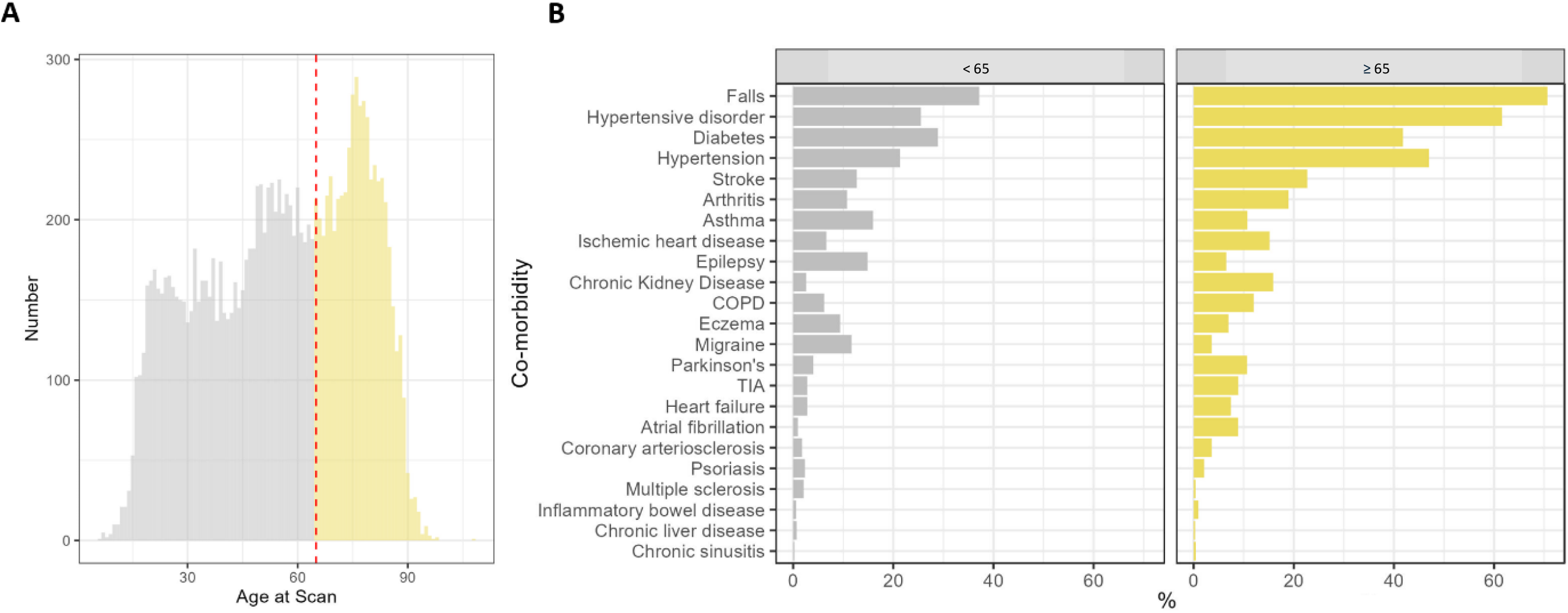
A) Histogram of the total number of MRI images acquired stratified by age with the red dashed line indicating a common cut off at 65 years old. B) Bar chart showing the percentage of comorbidities for linked MRI scans for patients <65 (left panel) and ≥ 65 years old (right panel)

On initial data curation, excluding unusable scans, we identified 14649 unique MR studies (exams) containing a total of 58620 scans. The total number of scans of each type is shown in Table 2.

**Table 2.**
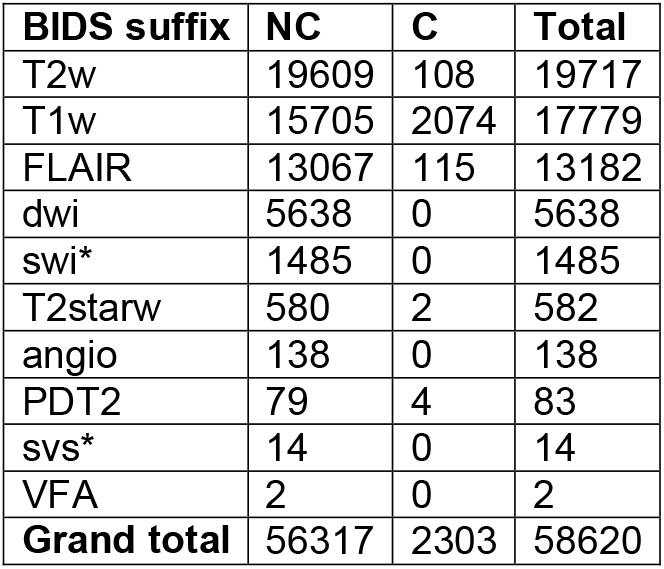
Total numbers of scans by BIDS suffix and contrast use.

Abbreviations (capitalisation as per BIDS): NC: non-contrast, C: contrast, T2w: T2-weighted, T1w: T1-weighted, FLAIR: fluid attenuated inversion recovery, dwi: diffusion weighted imaging, swi: susceptibility weighted imaging, T2starw: T2*-weighted, angio: angiogram, PDT2: dual echo PD and T2-weighted, svs: single voxel spectroscopy, VFA: variable flip angle, * indicates this suffix was in a draft or proposed version of BIDS at time of writing.

## Discussion

We have assembled an exceptionally rich large real-world cohort with linked MRI and clinical data reflecting the diversity of the population of South London that provides a unique platform for testing automated decision support tools. We hope that this cohort will provide clinically meaningful translation and application with an initial focus on identifying unique signatures of disease subtype and neuropsychiatric symptoms, markers of prognosis, and stratification for interventions and trials in a move towards real-world precision medicine. SLaM Image Bank leverages the strengths of CRIS to link brain MRI scans with clinical records.

Specific areas of interest relating to this cohort include using super-resolution approaches to improve image quality [14], automating volumetric and vascular analyses[31,32], normative modelling of disease cohorts with multimodal imaging data[33,34], and creating individual risk predictions that will be useful in trials and meaningful in clinical triage environments[35].

The use of real-world data has a number of strengths including socioeconomic and ethnic diversity with multiple co-morbidities and co-pathologies from people who are typically excluded from research cohorts. It is critical that AI models in neuroimaging are trained on these more representative datasets to address underlying biases, sources of disparity, and achieve diagnostic equity [1,12]. Another strength is that this is an imaging cohort naturally accruing from pre-existing data and does not require additional and expensive recruitment exercises. This is the first step that in the future may lead to a translational framework to build a powerful network of clinical sites with appropriate regulatory compliance. Furthermore, in contrast to highly selective research cohorts that concentrate on a single disease, our dataset comprises multiple diagnoses. This permits the development and/or validation of multi-class classifiers to distinguish between multiple diseases, or to develop transdiagnostic biomarkers.

Limitations of this cohort include the common limitations of real-world data from clinic such as the variable coding of routine clinical diagnoses and associated data, the variety and experience of healthcare raters, the likely greater co-morbidity than the general population, and the risk of under reporting variables that are not considered pathological or relevant to the underlying problem. Only MRI scans requested by SLaM are currently in this cohort and not other trusts within this geographical catchment. Mitigation strategies for this are the planned provision of linking outputs, training models and matching future imaging sequences in relation to normative population data such as from UK Biobank. Furthermore harmonisation of heterogenous clinical MRI scans to leverage research grade information using new AI tools applied to neuroimaging would facilitate this (13).

We feel that this new cohort provides a testbed for novel clinical decision support tools, provides unique epidemiological opportunities, and allows the exploration of novel real-world relationships with quantification of the relative weights of various factors on patient diagnosis, prognosis and stratification.

In the future we have the potential to add on various biomarkers to this database including other MRI modalities, routine CT and DAT, PET imaging, blood and CSF, based markers from neurodegeneration panels and linkage with other registries in the first instance. Further plans are to assess the impact of decision support tools on clinical management alongside assessing the health economic value and cost-effectiveness of such systems.

## Conclusion

We have assembled SLaM Image Bank as a rich real-world diverse London digital cohort linking MRI to clinical records for the development of clinical decision support tools using artificial intelligence and to provide novel epidemiological insights that have the potential to be clinically meaningful and translatable.

## Data Availability

The data are subject to the contractual restrictions of the data sharing agreements between South London and Maudsley NHS Trust and Kings College London and are therefore not available for access beyond the SLaM Image Bank research team without prior authorisation from the CRIS oversight (imaging) committee. Updates will be shared at www.brainregion.com/slamimagebank

## Acknowledgements

AVV is funded by the National Institute for Health Research (NIHR) as NIHR Clinical Lecturer and supported by the NIHR Maudsley Biomedical Research Centre and the NIHR HealthTech Research Centre in Brain Health at King’s College London and South London and Maudsley NHS Foundation Trust and King’s College London (NQOD-04) as PI of SLaM Image Bank. DS is part-funded by National Institute for Health Research (NIHR) Biomedical Research Centre at South London and Maudsley NHS Foundation Trust and King’s College London. RS is part-funded by: i) the National Institute for Health Research (NIHR) Biomedical Research Centre at the South London and Maudsley NHS Foundation Trust and King’s College London; ii) the National Institute for Health Research (NIHR) Applied Research Collaboration South London (NIHR ARC South London) at King’s College Hospital NHS Foundation Trust; iii) UKRI – Medical Research Council through the DATAMIND HDR UK Mental Health Data Hub (MRC reference: MR/W014386); iv) the UK Prevention Research Partnership (Violence, Health and Society; MR-VO49879/1), an initiative funded by UK Research and Innovation Councils, the Department of Health and Social Care (England) and the UK devolved administrations, and leading health research charities. The views expressed are those of the authors and not necessarily those of the NIHR or the Department of Health and Social Care.

